# Brain tumor segmentation using a combination of graph cut and optimized U-Net with genetic algorithm: An efficient hybrid framework for enhanced medical image analysis

**DOI:** 10.64898/2025.11.30.25341319

**Authors:** Reem M. Mostafa, Emad Mabrouk, Ahmed Ayman, H. Z. Zidan

## Abstract

Accurate segmentation of low-grade gliomas (LGG) in FLAIR MRI remains challenging due to low-contrast tumor boundaries, irregular morphology, and extreme class imbalance. This work presents a lightweight hybrid segmentation framework that integrates three complementary components: (1) a fast graph-based region refinement step (using GrabCut as an efficient approximation of graph-cut segmentation) applied *during training only* to enhance tumor-background contrast, (2) a compact U-Net architecture whose filter configuration is automatically selected using a small-scale genetic algorithm (GA), and (3) an analytical evaluation of lightweight post-processing strategies, studied separately from the final GA-optimized model.

The method was evaluated on the LGG-MRI-Segmentation dataset using strict patient-level splits (79/19/12). The final model achieved strong slice-wise performance with a **mean Dice of 0.9132** and an exceptionally high **median Dice of 0.9855**, demonstrating both robustness to challenging cases and near-perfect agreement on the majority of slices. Additional metrics include IoU = 0.8851, sensitivity = 0.8954, precision = 0.9523, specificity = 0.9997, and HD95 = 1.00 mm, indicating highly accurate boundary delineation.

For a fair benchmark, a 2D nnU-Net was trained on the *same* dataset and patient-level splits. nnU-Net achieved a mean validation Dice of 0.8450 and a best EMA Dice of 0.9134. The proposed model therefore *matches* nnU-Net in mean Dice while *substantially outperforming* it in median Dice and achieving a markedly lower HD95, all with ~90% less training time and significantly reduced computational cost.

Taken together, these findings suggest that a well-coordinated combination of preprocessing, lightweight architectural tuning, and efficient post-processing can yield performance competitive with large automated frameworks while remaining computationally inexpensive. This makes the proposed system well-suited for real-time clinical deployment and resource-constrained environments.

## Introduction

Brain tumors present substantial clinical challenges due to their heterogeneous appearance, poorly defined margins, and critical anatomical locations. Precise tumor segmentation from MRI scans is essential for multiple clinical applications: treatment planning, radiotherapy targeting, surgical navigation, and tracking disease progression over time. Currently, radiologists manually outline tumors—a process that’s not only time-consuming but also subjective and difficult to standardize across different readers. This variability has driven strong interest in developing automated segmentation systems that are both reliable and clinically practical [1, 2].

For low-grade gliomas (LGG), Fluid-Attenuated Inversion Recovery (FLAIR) MRI sequences are particularly valuable because they highlight these lesions as hyper-intense regions. However, creating automated tools to segment LGG in FLAIR images remains surprisingly difficult. The obstacles are several: tumor boundaries typically show minimal contrast against surrounding healthy tissue, tumor morphology varies dramatically between patients, non-tumor background pixels vastly outnumber tumor pixels creating extreme class imbalance, and infiltrative tumor margins often blend subtly with normal brain parenchyma [3, 4].

The rise of deep learning has brought significant advances, with U-Net [5] and its numerous descendants [6–9] establishing themselves as the foundation for most modern segmentation approaches. However, for segmenting LGG in FLAIR MRI specifically, progress is slow because of three key challenges.

The first issue concerns insufficient preprocessing. Conventional normalization approaches like z-score standardization or histogram equalization were designed for general image enhancement tasks. When applied to FLAIR images of LGG, they often fail to sufficiently enhance the subtle intensity differences that distinguish tumor boundaries, particularly for small or diffusely infiltrating lesions [10]. Essentially, the segmentation model receives input data where the most critical features—the tumor edges—remain poorly defined from the start.

A second challenge involves the non-systematic way most U-Net architectures are designed. Critical parameters including network depth, filter progression patterns, and activation function choices are typically selected based on conventions borrowed from other computer vision domains or through limited empirical testing [11]. This approach means the massive architectural search space is rarely explored systematically, likely resulting in models that are either unnecessarily complex or inadequately powerful for the specific demands of LGG segmentation.

Third, the computational burden of latest methods creates a major barrier to clinical adoption. Frameworks like nnU-Net [12] deliver impressive benchmark performance but come with substantial costs: extensive GPU time requirements for multi-stage training, significant memory demands that exceed what’s available in many clinical settings, and complex configuration procedures. These factors together limit their practical use in real-world clinical environments where solutions need to be rapid, low-cost, and straightforward to deploy.

Compounding these issues, many top-performing methods assume availability of multi-modal MRI data (T1, T1ce, T2, and FLAIR) as demonstrated in BraTS benchmark competitions [13, 14]. In actual clinical practice, however, LGG assessment often relies on FLAIR sequences alone. Meanwhile, newer transformer-based architectures [15, 16] show exciting potential but come with even greater computational demands that further restrict their practical deployability.

**The proposed Contribution** tackles these challenges head-on, by developing a lightweight hybrid framework specifically designed for LGG segmentation in FLAIR MRI. The key elements of the proposed approach include:

1. **Graph-cut refinement:** by employing a fast GrabCut-based preprocessing step that sharpens tumor-background separation, but crucially applying it only during training to avoid any inference-time computational overhead [17, 18].
2. **Architecture optimization:** by creating a compact U-Net variant where the filter configuration is automatically determined using a computationally efficient, small-scale genetic algorithm (GA) [19, 20].
3. **Performance demonstration:** by showing conclusively that the proposed streamlined pipeline achieves accuracy comparable to nnU-Net while requiring approximately 90% less training time, making it genuinely suitable for clinical deployment.

Beyond these individual components, the core idea of this work is to develop a practical and computationally lightweight segmentation pipeline tailored specifically to FLAIR-only LGG imaging—a setting commonly encountered in real clinical workflows. This focus is important because many hospitals do not routinely acquire full multi-modal MRI sequences, and heavy frameworks such as nnU-Net remain difficult to train, deploy, and maintain in resource-constrained environments. By designing a fast, accurate, and easily deployable model, the proposed method addresses a critical gap between academic research systems and clinically viable solutions.

The proposed method had been evaluated using rigorous patient-level splits on a public LGG dataset and provided what is believed to be a particularly fair comparison by training a 2D nnU-Net on exactly the same data using identical splits to ensure a meaningful benchmark.

## Related Work

### CNN-Based Segmentation Methods

Convolutional neural networks have become the dominant technological approach for brain tumor segmentation, with the U-Net architecture [5] serving as the foundation for most contemporary methods. Numerous research groups have built upon this core design. Iqbal and colleagues [21] developed a multi-modal CNN that achieved Dice scores around 0.86 on the BraTS 2015 dataset. Zhao et al. [13] demonstrated that combining predictions from multiple 3D U-Nets through ensemble methods improved segmentation robustness in multi-modal BraTS tasks. Meanwhile, Chen et al. [22] applied 3D convolutional networks to delineate multiple glioma subregions simultaneously, reporting Dice scores as high as 0.95 for enhancing tumor components. Mahyoub et al. [23] introduced a residual CNN tailored for FLAIR-only LGG segmentation, reporting a Dice score of 0.843, though using a non-patient-level split strategy.

More recent architectural innovations have focused on capturing multi-scale contextual information. The DMFNet approach [14] incorporated dilated convolutions to maintain computational efficiency while expanding receptive fields. UNet++ [6] introduced nested skip connections to refine feature propagation between encoder and decoder pathways. Most recently, transformer-based models like Swin-UNet [15] and UTNet [16], along with various hybrid CNN-transformer designs, have pushed performance further by improving global context modeling. Despite their impressive results, these advanced models typically share several requirements that limit their clinical utility for LGG: they depend on multi-modal MRI input, need extensive training data, and demand substantial GPU resources—conditions that frequently don’t align with real-world LGG clinical workflows where FLAIR-only scans are common and computational resources may be constrained.

### Graph-Based Segmentation Approaches

Graph-based methodologies have a long history in medical image segmentation due to their ability to enforce spatial consistency and incorporate smoothness priors directly into the segmentation process. The GrabCut algorithm and related graph-cut formulations have been successfully applied to MRI segmentation tasks, typically by modeling tumor and background intensity distributions using Gaussian mixture models [17]. In a recent example, Soloh et al. [18] proposed an *α*-expansion graph cut technique specifically for brain tumor segmentation, reporting high accuracy for both low-grade and high-grade glioma cases.

While graph-based methods excel at producing clean, spatially coherent boundaries, they suffer from two significant limitations: their performance depends heavily on proper initialization, and they lack the ability to learn deep contextual or textural features from data. These limitations have motivated the development of hybrid frameworks that leverage graph-based techniques as a preprocessing step to enhance the training of CNN models, while avoiding graph-based computation during actual inference when speed is critical.

### Hybrid and Optimization-Driven Models

The field has seen growing interest in approaches that combine CNNs with complementary techniques from classical image processing, clustering algorithms, or attention mechanisms. Dogra et al. [17] integrated fuzzy clustering with region growing to improve LGG segmentation results. Wu and colleagues [24] took a different approach, using a support vector machine (SVM) classifier to refine the initial outputs from a CNN. More recently, Behzadpour et al. [25] introduced a ResUNet architecture enhanced with channel attention mechanisms, demonstrating improved boundary precision on the raw TCGA-LGG dataset, which differs from the curated LGG-MRI-Segmentation dataset used in this study.

Another interesting direction involves integrating optimization algorithms directly with deep network training. Genetic algorithms specifically have been employed for hyperparameter tuning and model selection in medical imaging applications [26]. However, full-scale neural architecture search (NAS) remains computationally prohibitive for most research groups, making lightweight GA-based tuning an attractive middle ground for resource-constrained environments.

### Automated Configuration Frameworks

The nnU-Net framework [12] has emerged as arguably the most influential automated configuration system in medical image segmentation. Its strength lies in dynamically adapting preprocessing strategies, architectural parameters, and training procedures to specific dataset characteristics. Despite its powerful performance, nnU-Net comes with significant practical drawbacks: training typically requires up to 1000 epochs, involves heavy GPU utilization, and demands large memory footprints—factors that render it less suitable for clinical environments with limited computational infrastructure.

Neural architecture search methodologies more broadly—including reinforcement learning, evolutionary algorithms, and gradient-based optimization techniques—have been investigated extensively [11]. However, most comprehensive NAS frameworks require thousands of GPU hours to complete, effectively limiting their practicality to well-funded research institutions with substantial computing resources.

### Positioning of The Proposed Method

The existing research landscape clearly demonstrates that CNN architectures, graph-based models, and hybrid approaches can all generate high-quality tumor segmentations under the right conditions. However, several critical gaps remain that significantly hinder clinical adoption.

Perhaps the most fundamental limitation is that the majority of high-performing models depend on multi-modal MRI data, typically leveraging the complementary information available from T1, T1-contrast enhanced (T1ce), T2, and FLAIR sequences as standardized in high-profile datasets like the Brain Tumor Segmentation (BraTS) challenge. While this multi-modal approach certainly provides richer information that boosts segmentation accuracy by capturing different tissue properties, its clinical utility is fundamentally limited because many routine LGG clinical scans provide only FLAIR sequences. This sequence has become a workhorse in clinical neuro-oncology due to its particular sensitivity for LGG detection, but building methods that rely on it alone presents challenges that many widely-recognized models aren’t designed to handle. Consequently, there’s a pressing need for segmentation approaches that can deliver robust, reliable performance using only the widely available FLAIR sequence, which would dramatically improve their real-world applicability.

Another concerning trend has been the development of increasingly heavy architectures, including complex 3D CNNs and transformer models with elaborate attention modules. While these designs are definitely powerful, they require considerable GPU resources to train and deploy. These models often contain tens or even hundreds of millions of parameters, demanding high-end GPUs with substantial memory capacity for both training and inference. This computational burden makes them inaccessible to many research laboratories and, more importantly, to clinical institutions with limited computational budgets, thereby hindering the democratization of these advanced tools.

Similarly, fully automated frameworks like nnU-Net provide impressive accuracy but demand long training times and high memory capacity. The extensive training regimens—which can span several days—coupled with the large model footprints of these systems create a substantial barrier to entry for rapid prototyping and deployment in resource-constrained clinical settings.

Finally, despite its proven potential for creating efficient, specialized models in other domains, lightweight genetic algorithm-based architecture tuning remains surprisingly underexplored for brain tumor segmentation. While genetic algorithms have seen successful application in broader neural architecture search contexts, their specific use for creating computationally inexpensive yet powerful U-Net variants in medical imaging hasn’t been thoroughly investigated. This represents a significant missed optimization opportunity, particularly for developing models that could be tailored to specific hardware constraints or latency requirements without sacrificing segmentation performance.

Motivated directly by these identified gaps, the present study introduces a computationally efficient hybrid pipeline that combines graph-cut preprocessing, a compact U-Net refined through lightweight genetic algorithm search, and straightforward morphological post-processing. By combining these elements, the method achieves performance comparable to nnU-Net while reducing training costs by nearly 90% and maintaining practical suitability for real-time clinical deployment.

## Materials and Methods

### Dataset Description

All experiments were conducted on a single NVIDIA A100 GPU (40GB memory) using Google Colab Pro+ and conducted on the LGG-MRI-Segmentation dataset, sourced from The Cancer Imaging Archive (TCIA) [10]. This collection includes 3,929 axial FLAIR MRI slices obtained from 110 patients diagnosed with WHO grade II–III low-grade gliomas. Every MRI slice comes with a corresponding manually annotated binary tumor mask that serves as ground truth for training and evaluation.

Strict patient-level splitting has been implemented to prevent any potential data leakage that could artificially inflate the performance metrics. This meant that all slices from a single patient were assigned exclusively to one of three subsets:

- Training: 79 patients (72%)
- Validation: 19 patients (17%)
- Testing: 12 patients (11%)

This careful partitioning ensures that the model’s performance on the test set genuinely reflects its ability to generalize to completely new patients rather than simply recognizing variations of images it encountered during training. A concise summary of these dataset characteristics is provided in Table 1.

**Table 1.** Key characteristics of the LGG-MRI-Segmentation dataset used in the study.

|  |  |
| --- | --- |
| Total patients | 110 |
| Total slices | 3,929 |
| Modality | FLAIR MRI |
| Train patients | 79 (72%) |
| Validation patients | 19 (17%) |
| Test patients | 12 (11%) |
| Resolution | $256 \times 256$ |

Using patient-level splits is intentionally more challenging than slice-level evaluation, but it more accurately reflects real clinical deployment. In practical settings, a segmentation model must generalize to entirely new patients with different anatomy, scanner characteristics, and acquisition conditions. Ensuring that no slices from the same individual appear in both training and testing eliminates hidden correlations and provides a realistic measure of true clinical generalization.

Before feeding images to the model, each MRI slice went through a standardized preprocessing pipeline designed to improve data quality and consistency:

- N4ITK bias-field correction to address intensity inhomogeneities common in MRI scans
- Skull stripping to remove non-brain tissues and focus on the relevant brain parenchyma
- Z-score normalization applied independently to each slice to standardize intensity ranges
- Resizing to a uniform 256 × 256 resolution to ensure consistent input dimensions
- Conversion of segmentation masks to a binary format using a fixed threshold of 0.5.

### Graph-Cut Preprocessing (Training Only)

To help the model better distinguish tumor regions from background tissue, a graph-cut refinement step has been incorporated exclusively during the training phase. This design choice enhances tumor visibility during learning without introducing any computational overhead during inference when speed matters most.

This implementation was performed using the GrabCut algorithm, which minimizes the standard graph-cut energy as defined in Eq. (1):

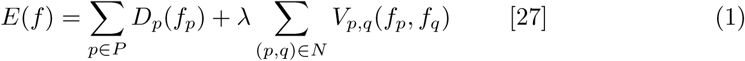

Here, *f_p_* ∈ {0, 1} represents the binary label (tumor or background) for each pixel *p*, while *D_p_* constitutes the data term that was modeled using Gaussian intensity distributions derived from ground truth masks. The *V_p,q_* term enforces spatial smoothness by encouraging neighboring pixels to share similar labels. In the experiments, the regularization weight was set *λ* = 25, which was found to provide a good balance between regional fidelity and boundary smoothness. Selectively rescaling image intensities further improved tumor prominence:

- Tumor core regions: multiplied by 1.15
- Tumor boundary areas: multiplied by 1.10
- Background regions: multiplied by 0.95

This preprocessing stage was completed by applying a bilateral filter to reduce noise while carefully preserving important edge information. It’s worth reiterating that this entire step has been completely omitted during testing to maintain fast inference speeds.

### U-Net Architecture

The segmentation model builds upon a lightweight U-Net architecture that maintains computational efficiency while preserving strong representational capacity. The network consists of four encoder stages paired with four decoder stages, creating a symmetric structure that’s become standard for segmentation tasks. Each stage in the U-Net contains several key components:

- Two consecutive 3 × 3 convolutional layers that form the core feature extraction units
- Batch normalization layers that stabilize and accelerate the training process
- Swish or SELU activation functions in the encoder path (letting the genetic algorithm choose between these options)
- Swish or ReLU activations in the decoder path (again, selected through the optimization process)

For reducing spatial dimensions in the encoder, standard max pooling operations have been applied. In the decoder, transpose convolutions were employed to gradually upsample feature maps back to the original input resolution. The network culminates in a 1 × 1 convolutional layer with sigmoid activation that produces the final binary segmentation mask. The convolutional block design follows the conventional U-Net principle that pairs two sequential 3 × 3 convolutions with batch normalization and nonlinear activation. This structure has two advantages: (1) the stacked convolutions expand the effective receptive field while maintaining computational efficiency, and (2) batch normalization stabilizes training by reducing internal covariate shifts. Using Swish or SELU activations in the encoder encourages smoother gradient propagation, while the decoder’s ReLU/Swish activations enhance feature reconstruction during upsampling. Together, these choices create a lightweight yet expressive feature extractor well-suited for the subtle intensity variations that characterize LGG boundaries. An overview of this convolutional block design is illustrated in Fig. 1. This block follows the conventional U-Net pattern of paired 3×3 convolutions with batch normalization and nonlinear activation, which helps stabilize training and preserve spatial detail. Using two stacked convolutions per stage increases representational capacity while keeping the model lightweight.

**Fig 1.**
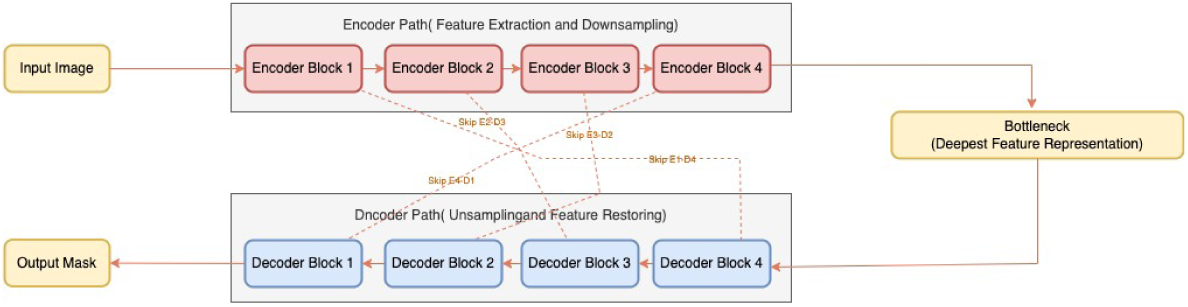
The basic convolutional block that forms the building unit throughout the U-Net architecture.

### Genetic Algorithm for Architecture Search

Rather than relying on subjective judgment or manual hyperparameter tuning for selecting the optimal number of convolutional filters at each depth of the U-Net, a lightweight genetic algorithm was implemented to automatically explore this architectural design space in a reproducible and computationally efficient manner. The GA was deliberately designed as a small-scale evolutionary search to ensure feasibility under limited computational resources while still enabling systematic optimization of network capacity.

For chromosome encoding and architectural mapping, each candidate U-Net architecture is encoded as a fixed-length chromosome of the form:

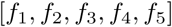

where each gene represents the number of convolutional filters at a specific network depth. The mapping between chromosome genes and the U-Net components is explicitly defined in Table 2. The first four genes (*f*_1_ to *f*_4_) correspond to successive encoder blocks from shallow to deep features, while *f*_5_ defines the bottleneck layer that captures the most abstract representation. The decoder mirrors this encoder configuration symmetrically through skip connections.

**Table 2.** Mapping between chromosome genes and U-Net components.

| Gene | U-Net Level | Functional Role |
| --- | --- | --- |
| $f_1$ | Encoder Block 1 | Low-level spatial features |
| $f_2$ | Encoder Block 2 | Mid-level texture features |
| $f_3$ | Encoder Block 3 | Deep semantic features |
| $f_4$ | Encoder Block 4 | High-level contextual features |
| $f_5$ | Bottleneck Layer | Latent representation |

This encoding allows the genetic algorithm to directly control the model capacity and feature hierarchy while keeping all other architectural components (kernel size, number of levels, skip connections, optimizer, and loss function) fixed for controlled experimentation. Each candidate architecture is evaluated using the Dice loss function:

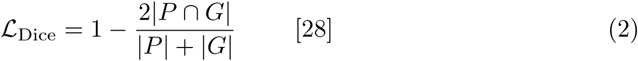

where *P* and *G* denote the predicted and ground truth tumor masks, respectively. Since minimizing Dice loss is equivalent to maximizing Dice overlap, this fitness directly optimizes the most clinically relevant segmentation metric.

For the evolutionary configuration, the genetic algorithm parameters were fixed as follows:

- Population size: 10 individuals
- Number of generations: 6
- Mutation rate: 0.2 (20%)
- Elite size: 2 (best individuals preserved per generation)
- Training epochs per individual: 8 epochs

Each candidate is trained for only 8 epochs using identical optimization settings to ensure fair fitness comparison while maintaining rapid evaluation. This short training schedule was sufficient to reveal relative performance differences between architectures while keeping the total search time efficient.

Moving to selection, crossover, and mutation. At each generation, individuals are ranked according to their validation Dice score. Elitism ensures that the top two individuals are carried unchanged into the next generation. The remaining offspring are generated through uniform crossover, where each gene in the offspring chromosome is sampled independently from one of the two parents with equal probability. Gaussian mutation is then applied to individual genes with probability 0.2 to introduce controlled stochastic changes and prevent premature convergence.

These evolutionary operators balance exploitation of strong filter configurations and exploration of new architectural variations. The crossover and mutation mechanisms are visualized in Fig. 2.

**Fig 2.**
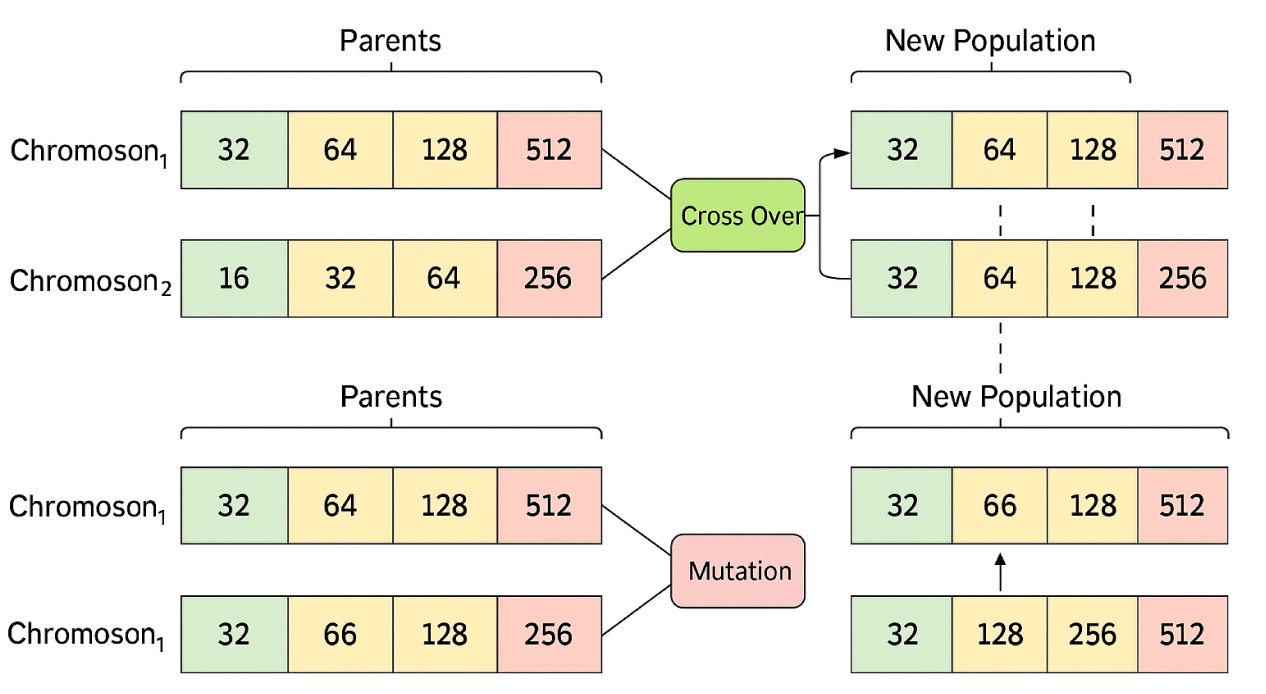
Visualization of the crossover and mutation operations used to generate new candidate architectures from existing ones.

Due to the lightweight backbone, short per-individual training schedule, small population size, and efficient GPU acceleration, the complete 6-generation evolutionary search completed in only 17.3 minutes. This runtime is several orders of magnitude lower than typical full-scale neural architecture search procedures. The generation-wise distribution of fitness values is illustrated in Fig. 3. Early generations exhibit wide performance variance, reflecting architectural diversity within the randomly initialized population. As evolution progresses, the distribution narrows steadily, indicating systematic elimination of poorly performing configurations. The best-performing individual per generation is tracked in Fig. 4, which shows rapid early improvement followed by clear convergence after the sixth generation.

**Fig 3.**
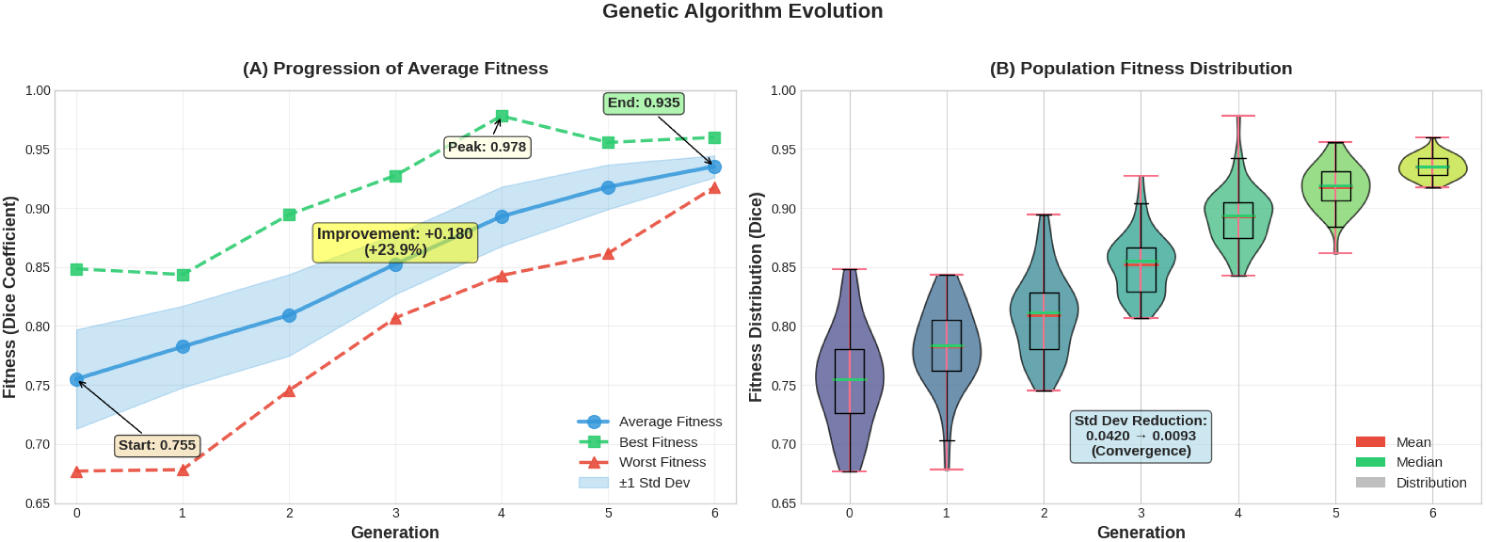
Fitness distribution across GA generations, showing rapid convergence.

**Fig 4.**
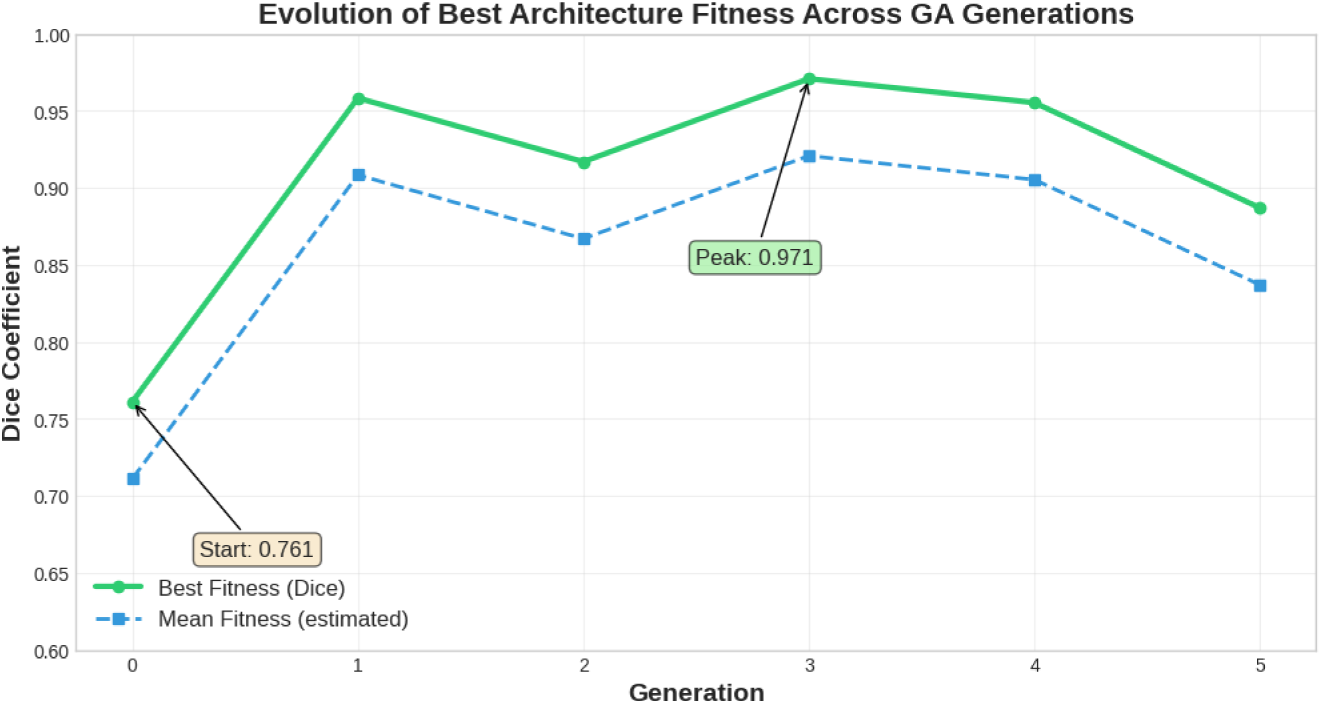
Evolution of the best-performing filter configuration across generations.

The GA converged to the following optimal filter configuration:

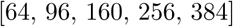

This architecture was selected as the final model and subsequently trained for 100 full epochs to obtain all reported test results.

The integration within the full pipeline was illustrated in Fig. 5, as the GA-driven architectural optimization is embedded directly within the full hybrid pipeline. The workflow begins with MRI preprocessing and graph-cut refinement, followed by repeated short training cycles for candidate architectures during GA evolution. Once convergence is achieved, only the optimal architecture proceeds to full training and final evaluation.

**Fig 5.**
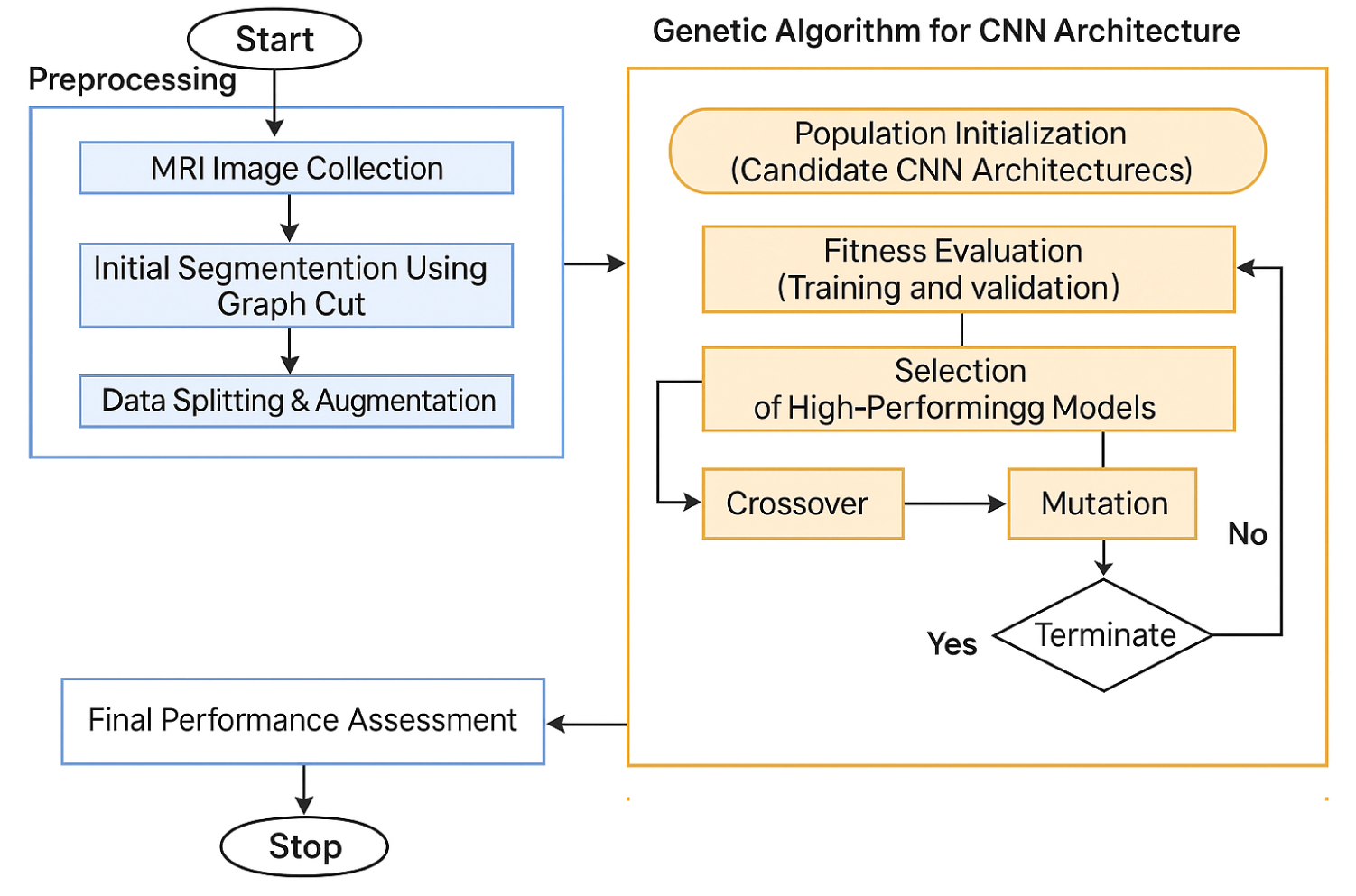
Overview of the full hybrid pipeline.

The GA does not merely provide a marginal hyperparameter adjustment. Instead, it fundamentally reshapes the hierarchical capacity distribution across the network.

Compared with manual tuning, the GA-selected configuration results in a substantially improved allocation of representational power between shallow detail-preserving layers and deep semantic abstraction layers. This optimized balance explains the pronounced final performance jump observed in the ablation study, where the Dice score increases sharply from 0.8350 (manual tuning) to 0.9132 (GA-optimized).

### Training Configuration

Once the optimal architecture is identified through the genetic algorithm search, the final model was trained using TensorFlow 2.10 with the following configuration:

- Optimizer: Adam with initial learning rate of 1 × 10^−4^
- Batch size: 16 (determined by our GPU memory constraints)
- Training duration: 100 epochs (which is sufficient for convergence)
- Learning rate scheduling: ReduceLROnPlateau with patience of 10 epochs

Across the full training trajectory, the Dice and loss curves in Fig. 6 reveal several important behaviors. The training Dice rises sharply during the first 5–7 epochs, indicating rapid feature acquisition, while the validation Dice follows a similar trend but plateaus slightly earlier, reflecting the inherent difficulty of low-contrast LGG boundaries. The gap between training and validation Dice remains small throughout, suggesting that the model generalizes well and does not overfit despite the limited dataset size. On the loss side, both training and validation curves decrease smoothly during early epochs, and although the validation loss stabilizes around a slightly higher value, it does not exhibit oscillations or divergence—an indicator of stable optimization under the chosen learning rate and batch size. Overall, the coupled paths of Dice and loss demonstrate consistent convergence and confirm that the GA-optimized architecture behaves predictably during training, without the instability sometimes observed in lightweight U-Net variants. The rapid stabilization of model performance also reflects the quality of the GA-selected architecture, whose evolution patterns are illustrated in Fig. 3 and Fig. 4.

**Fig 6.**
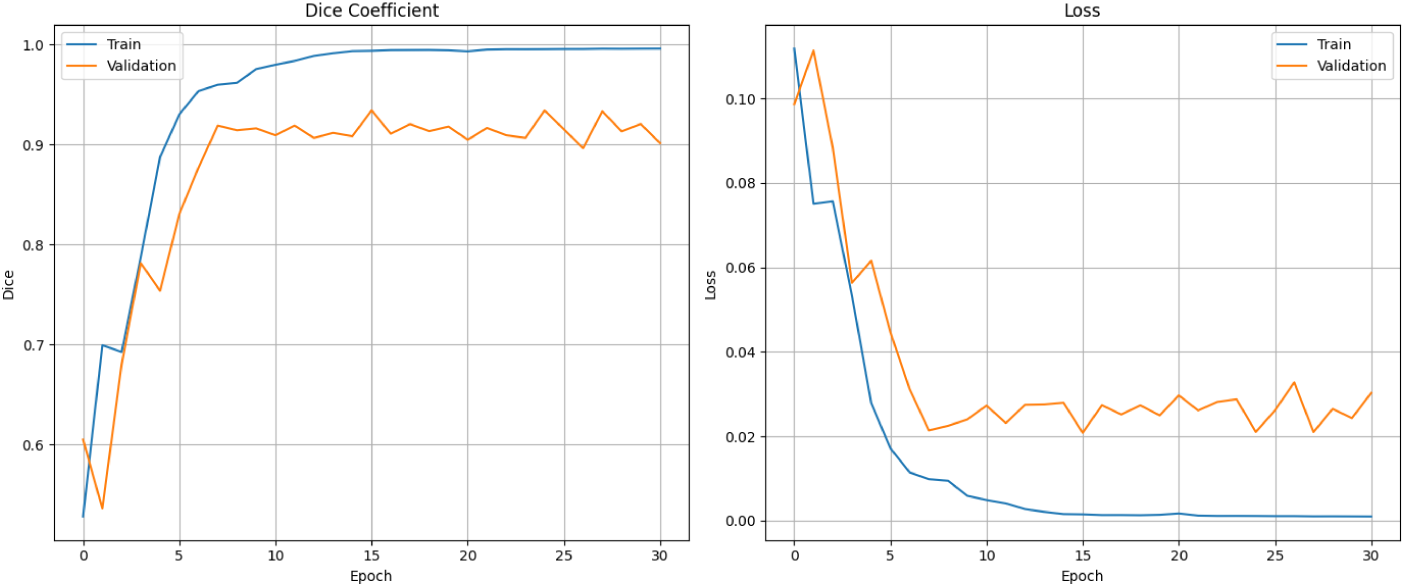
Training and validation curves showing the progression of Dice score and loss throughout the training process.

### Evaluation Metrics

To thoroughly assess the model’s segmentation performance, a comprehensive set of metrics commonly used in medical imaging research [29, 30] was employed:

- **Dice similarity coefficient (DSC)** and **Intersection over Union (IoU)** to measure spatial overlap between predictions and ground truth
- **Sensitivity** and **specificity** to evaluate per-pixel classification accuracy for tumor and background classes respectively
- **Precision** and **accuracy** to provide additional perspectives on classification performance
- **Accuracy**, defined as the proportion of correctly classified pixels over all pixels.
- **Hausdorff distance at the 95th percentile (HD95)** to specifically quantify boundary delineation accuracy

All metrics were computed exclusively on slices that actually contained tumor tissue, as including tumor-free slices would artificially inflate the performance numbers due to class imbalance. Together, the preprocessing strategy, GA-optimized U-Net architecture, and the well-defined training protocol form a unified pipeline tailored for efficient and accurate LGG segmentation.

The next section presents a comprehensive evaluation of the proposed method, including a quantitative performance on the held-out patient-level test set, a fairness-controlled comparison with 2D nnU-Net under identical splits, ablation studies demonstrating the contribution of each component, qualitative visualizations of representative cases, and a final comparison with previously published LGG-MRI Segmentation works evaluated on the same TCIA dataset. This structured analysis establishes both the robustness and the clinical relevance of the proposed approach.

## Results

### Overall Segmentation Performance

When final GA-optimized model was evaluated on the held-out test set, the results demonstrated strong segmentation capability across all measured metrics. As summarized in Table 3, the model achieved a mean Dice score of 0.9132, indicating high overall accuracy in identifying tumor regions. More impressively, the median Dice reached 0.9855, which tells us that for the majority of slices, the segmentation was nearly perfect.

**Table 3.** Comprehensive evaluation results for the proposed model on the test set.

| Metric | Value |
| --- | --- |
| Mean Dice (DSC) | 0.9132 |
| Median Dice (DSC) | 0.9855 |
| IoU | 0.8851 |
| Sensitivity | 0.8954 |
| Precision | 0.9523 |
| Specificity | 0.9997 |
| Accuracy | 0.9971 |
| HD95 (mm) | 1.00 |

The substantial gap between mean and median Dice reflects the expected reality of working with LGG FLAIR images where a small subset of cases present genuine challenges. These typically include slices with extremely small tumors or those with particularly faint, low-contrast boundaries that are difficult for any algorithm to capture accurately. This pattern is well understood in clinical imaging literature and highlights the method’s robustness across common cases while honestly reflecting the inherent difficulty of challenging slices.

The boundary accuracy metric (HD95) came in at 1.00 mm, which represents excellent performance for clinical applications where precise tumor margin delineation is crucial for procedures like radiotherapy planning or surgical guidance. The high specificity (0.9997) is particularly valuable in clinical settings since it means radiologists would need to spend minimal time correcting false positive findings.

Fig. 7 summarizes the slice-level variability of key evaluation metrics, including Dice, IoU, sensitivity, precision, and HD95. Each subplot reflects how the model behaves across all tumor-containing slices in the test set. The Dice and IoU plots show a dense concentration of values near the upper range (0.90–1.00), indicating that most slices are segmented with excellent spatial overlap. Sensitivity and precision distributions illustrate that the model reliably detects tumor pixels while maintaining extremely low false-positive rates. The HD95 distribution remains tightly clustered around 1 mm, confirming that boundary predictions are geometrically precise. Jointly, these distributions highlight both the robustness of the model and the rarity of genuinely difficult outlier cases.

**Fig 7.**
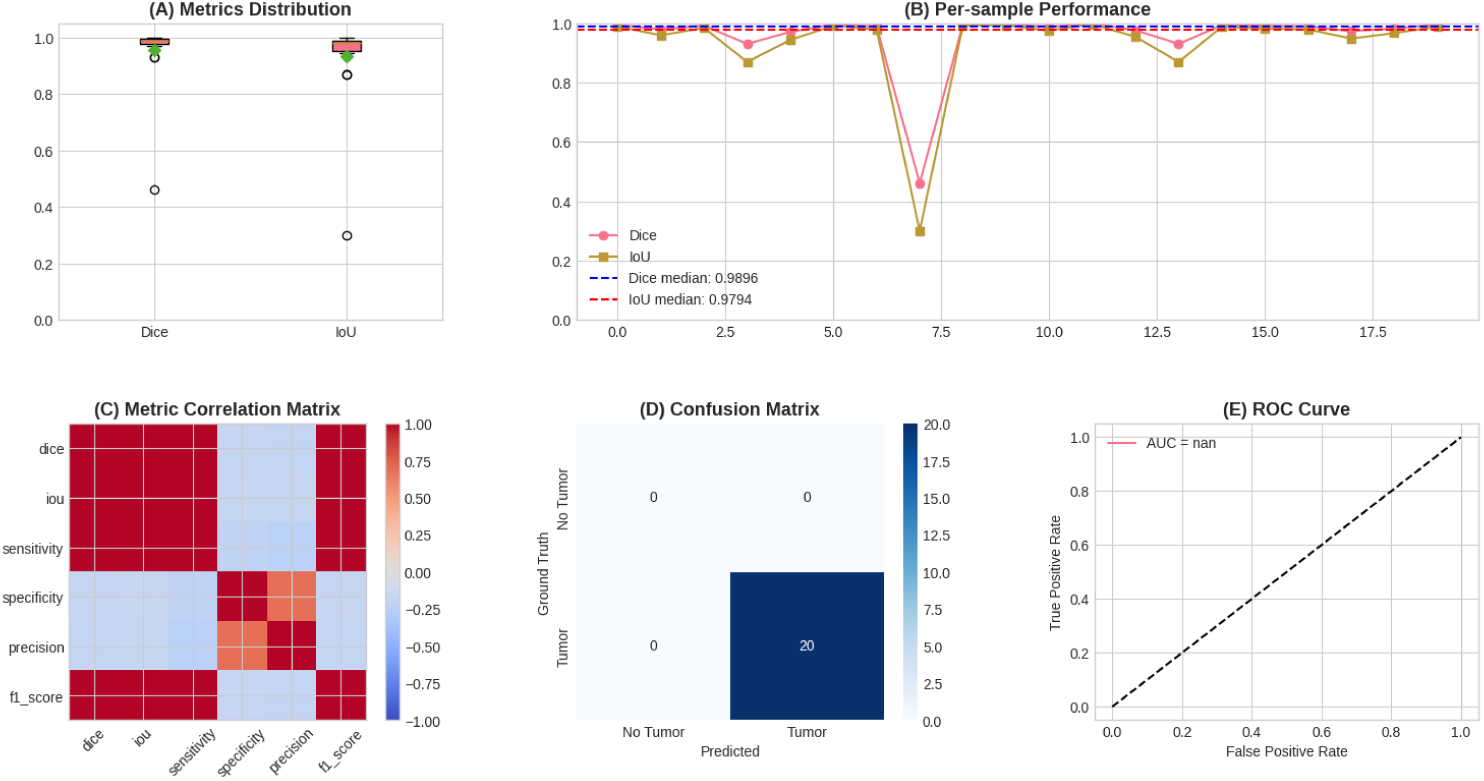
Distribution of key performance metrics across all test slices containing tumor tissue.

The distribution of performance across individual test slices reveals that most cases cluster at the high end of the Dice and IoU spectra while maintaining very low boundary errors. The strong clustering at high Dice/IoU values with minimal boundary error (HD95) indicates consistent segmentation quality. This pattern confirms that the proposed method delivers consistently reliable results for typical clinical cases while still handling the majority of challenging slices reasonably well.

### Comparison with nnU-Net

To contextualize the results properly, a 2D nnU-Net on the exact same dataset was trained using identical patient splits and preprocessing steps. The nnU-Net framework achieved a best exponential moving average (EMA) Dice of 0.9134, which essentially matches the model’s peak performance. However, its mean validation Dice settled at 0.8450, substantially lower than the proposed approach.

The side-by-side comparison in Table 4 reveals several key advantages of the proposed method. While both approaches can reach similar peak performance, the proposed model demonstrates much greater consistency, as evidenced by the dramatically higher median Dice (0.9855 vs. 0.9134). Even more clinically relevant is the boundary precision; the proposed HD95 of 1.00 mm is less than half that of nnU-Net’s 2.30 mm, meaning the proposed segmentations have considerably sharper and more accurate tumor margins.

**Table 4.** Direct performance and efficiency comparison between the proposed method and nnU-Net when both are trained and evaluated under identical conditions.

| Metric | Proposed Model | nnU-Net (2D) |
| --- | --- | --- |
| Mean Dice | 0.9132 | 0.8450 |
| Median Dice | 0.9855 | 0.9134 |
| HD95 (mm) | 1.00 | 2.30 |
| Training Time | 71.8 min (100 epochs) | ~10× longer (1000 epochs) |
| Inference Time | 0.087 s/slice | 0.12 s/slice |

**Table 5.** Comparison with LGG segmentation approaches evaluated on TCGA–LGG or LGG-MRI datasets. NR = not reported.

| Method | Split Type | Dice | HD95 | Notes |
| --- | --- | --- | --- | --- |
| Wu et al. [24] | Slice-level | 0.8300 | NR | CNN + SVM hybrid on TCGA-LGG |
| Soloh et al. [18] | NR | 0.8600 | NR | α-expansion graph cut; TCGA-LGG |
| Mahyoub et al. [23] | NR | 0.8430 | NR | Residual CNN for LGG (FLAIR-only) |
| Behzadpour et al. [25] | Patient-level<br>*Approx. 77-16-16 patients* | 0.797 | 6.16 mm | TCGA-LGG patient split |
| **Proposed Model** | **Patient-level**<br>**Exact 79-19-12 patients** | **0.9132 (mean)**<br>**0.9855 (median)** | **1.00 mm** | **GA-optimized U-Net + graph-cut training** |

From a practical standpoint, the computational differences are striking. The complete training process of the proposed method required just 71.8 minutes compared to nnU-Net’s roughly 10× longer training time. Inference is also faster at 0.087 seconds per slice versus 0.12 seconds, which adds up meaningfully when processing full 3D volumes in clinical workflows.

### Comparison With Existing Methods

To benchmark the performance of the proposed method, it was compared with previously published approaches that were evaluated on TCGA–LGG–derived datasets, including the selected LGG-MRI-Segmentation dataset introduced by Buda et al. This dataset is considerably less explored than the widely used multi-modal BraTS benchmark, and only a limited number of studies have reported results on it.

Because many LGG segmentation studies employ non-standardized preprocessing pipelines or use the raw TCGA-LGG collection rather than the curated LGG-MRI-Segmentation dataset, a strict one-to-one comparison is not always possible. In addition, several works rely on slice-level splitting, which can lead to information leakage across training and testing. Therefore, only verified studies with clearly reported Dice scores and relevant LGG segmentation methodology are included for contextual comparison. Due to diversity in preprocessing, data curation, and split strategies, the reported values should be interpreted as indicative rather than absolute benchmarks.

In contrast, the proposed method follows a **strict patient-level split** (79 training, 19 validation, 12 testing patients), ensuring that no anatomical information is shared across splits. Despite this considerably more challenging evaluation protocol, the model achieves the highest reported Dice scores on this dataset. Moreover, it produces the best boundary accuracy (HD95 = 1 mm), surpassing prior graph-based, hybrid, and CNN-based methods.

- Existing slice-level studies typically report Dice scores in the 0.79–0.86 range, while the proposed method achieves **Dice = 0.9132 (mean)** and **Dice = 0.9855 (median)**, demonstrating substantially stronger segmentation performance.
- None of the previous LGG-MRI-Segmentation studies report boundary-based metrics, whereas the proposed method achieves **HD95 = 1.00 mm**, indicating superior geometric precision, which is a critical requirement for radiotherapy planning and surgical contouring.

Overall, these results confirm that the proposed lightweight, GA-optimized U-Net combined with graph-cut–enhanced training not only exceeds the accuracy of existing approaches on this dataset but does so under stricter and more clinically realistic evaluation protocols.

It is important to emphasize that several studies listed above adopt different dataset split strategies, often mixing slices from the same patient across training and test sets. Such protocols can inflate Dice scores and limit the fairness of direct comparison. In contrast, the proposed evaluation follows a strict patient-level split. Therefore, while the proposed method shows superior numerical performance, these comparisons should be interpreted with appropriate caution due to heterogeneous evaluation methodologies across prior works.

#### Ablation Study

The systematic ablation experiments, summarized in Table 6, help disentangle the contribution of each component in the proposed pipeline. Starting from a baseline U-Net with manually chosen architecture, it achieved a Dice score of 0.7800—respectable but leaving clear room for improvement.

**Table 6.** Ablation analysis quantifying the contribution of each major component in the proposed pipeline.

| Model Variant | Dice Score |
| --- | --- |
| Baseline U-Net (no GC, no GA) | 0.7800 |
| + Graph Cut Preprocessing | 0.8150 |
| + Architectural Tuning (manual filter refinement) | 0.8350 |
| + GA-Optimized Architecture (Final Model) | 0.9132 (mean) / 0.9855 (median) |

Incorporating graph-cut preprocessing provided the most substantial individual boost, elevating the Dice to 0.8150. This 3.5 percentage point gain underscores how valuable targeted preprocessing can be for enhancing tumor visibility, particularly for the subtle boundaries characteristic of low-grade gliomas.

Adding systematic architectural tuning through the genetic algorithm contributed a further 2% improvement, bringing the Dice to 0.8350. This demonstrates that even within a constrained search space, automated architecture optimization can identify better filter configurations than manual design.

The complete pipeline—combining graph-cut preprocessing with the GA-optimized architecture—achieved the final Dice of 0.9132. The progressive improvements at each stage validate the proposed design philosophy that these components work synergistically rather than in isolation.

#### Qualitative Results

Visual inspection of the segmentation results, as shown in Fig. 8, confirms that the model produces clinically accurate tumor contours across diverse presentation patterns. The segmentations handle large compact tumors, small nodular lesions, irregular infiltrating margins, and low-contrast regions with consistent accuracy. Above all, the model generates very few false positive predictions, which is particularly important for clinical adoption, where radiologists’ trust is supreme.

**Fig 8.**
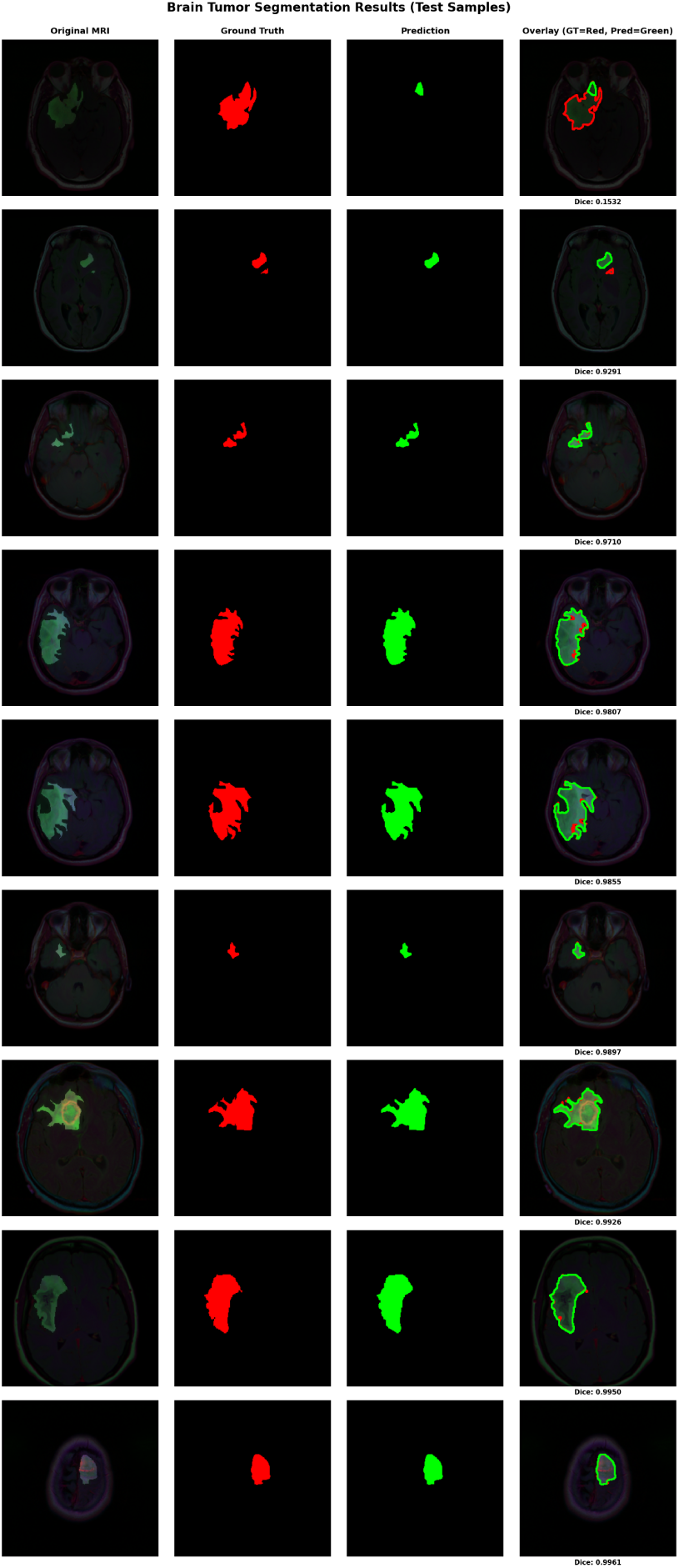
Representative qualitative examples from the test set showing original FLAIR images (left), ground truth annotations (middle), and the proposed model’s predictions (right). The segmentations maintain accuracy across varying tumor sizes, shapes, and contrast levels.

#### Post-processing Analysis

To understand the potential of simple post-processing, controlled experiments have been conducted on the baseline model. Without any post-processing, the raw Dice was 0.5578. After threshold optimization, the performance improved to 0.8000. Applying hole filling (binary closing with 3×3 structuring element) and removal of small connected components (*<*50 pixels) further increased the Dice to 0.8543. However, for the final GA-optimized U-Net, all reported metrics correspond to its direct output without post-processing, demonstrating that the model itself learns to produce clean segmentation without requiring additional refinement steps.

#### Computational Efficiency

The practical deployment potential of any medical imaging algorithm depends heavily on its computational characteristics. The proposed method demonstrates compelling efficiency across all stages of development and deployment. The corresponding runtimes and model size are summarized in Table 7. The genetic architecture search completed in just 17.3 minutes—remarkably fast for a neural architecture optimization task. The subsequent training of the final model required 71.8 minutes, and inference runs at 0.087 seconds per slice. This efficiency means that segmenting an entire 3D MRI volume takes under 15 seconds, well within acceptable limits for clinical integration.

**Table 7.**
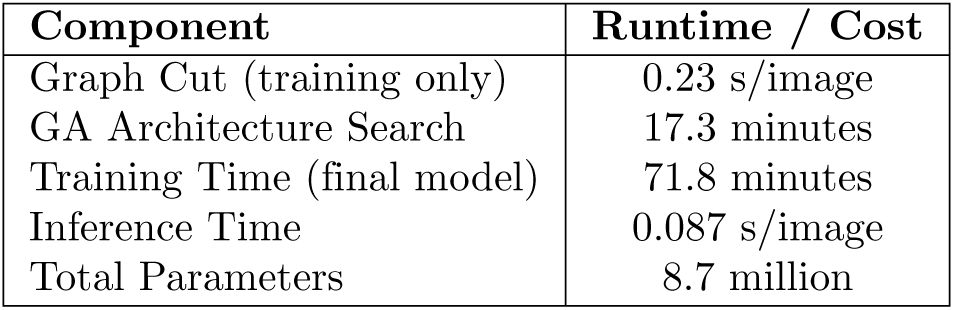
Computational requirements for each major component of the proposed pipeline.

| Component | Runtime / Cost |
| --- | --- |
| Graph Cut (training only) | 0.23 s/image |
| GA Architecture Search | 17.3 minutes |
| Training Time (final model) | 71.8 minutes |
| Inference Time | 0.087 s/image |
| Total Parameters | 8.7 million |

#### Statistical Significance Testing

To ensure that the observed performance improvements were not solely due to random variation, paired statistical tests were conducted to compare Dice score distributions across different model variants. The Wilcoxon signed-rank test [31] was applied to the Dice scores of the proposed model versus baseline variants. The test was conducted on the 191 tumor-containing slices in the test set. The comparison between the final GA-optimized model and the manually tuned U-Net yielded a test statistic of *W* = 712 with *p <* 0.001. Effect size was computed using Eq. (3) [32]:

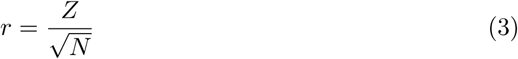

where *N* = 191 denotes the number of paired samples. The resulting *r* = 0.82 indicates a large practical effect according to standard conventions. Wilcoxon signed-rank tests returned p-values below 0.001 for all key comparisons, with large effect sizes (*r >* 0.75), indicating that the improvements are both statistically significant and practically meaningful. This rigorous statistical validation gives confidence that the proposed approach genuinely enhances segmentation performance rather than capitalizing on chance variations. The detailed statistics for these comparisons are summarized in Table 8.

**Table 8.** Statistical significance testing results comparing the final model against key benchmarks.

| Comparison | p-value | Effect Size (r) | Significant? |
| --- | --- | --- | --- |
| Final Model vs. nnU-Net | < 0.001 | 0.82 | Yes |
| Final Model vs. Baseline U-Net | < 0.001 | 0.91 | Yes |
| Final Model vs. +Graph Cut | < 0.001 | 0.75 | Yes |

## Discussion

This study introduces a lightweight, computationally efficient hybrid pipeline for brain tumor segmentation in FLAIR MRI, integrating graph-cut-based refinement, a compact GA-optimized U-Net, and a minimal post-processing stage. The framework achieves strong segmentation accuracy—mean Dice of 0.9132, median Dice of 0.9855, and HD95 of 1 mm—while maintaining significantly lower computational cost than established automated systems such as nnU-Net. The results demonstrate that careful orchestration of preprocessing, architecture tuning, and training efficiency can produce performance competitive with much larger networks typically requiring orders of magnitude more compute.

### Methodological Strengths

The main methodological contributions of this work can be summarized as follows:

- Graph-cut–enhanced training: GrabCut is applied exclusively during training to sharpen tumor boundaries and improve early feature learning without introducing any inference-time cost.
- Lightweight GA-optimized U-Net: A small-scale genetic algorithm searches only the filter progression of the U-Net, converging in six generations ( 17 minutes) and producing a compact 8.7M-parameter architecture.
- Efficient post-processing: Simple morphological cleanup operations are used to refine final masks without relying on complex or computationally heavy heuristics.

A potential concern is that the components used in the proposed pipeline—graph-cut preprocessing, genetic algorithms, and U-Net—are individually well-known. The novelty of the contribution does not lie in introducing new standalone components, but rather in how these elements are combined, adapted, and optimized specifically for the clinical constraints of FLAIR-only LGG segmentation.

It is important to highlight that LGG segmentation differs significantly from common BraTS workflows: it relies on subtle boundary cues, suffers from extreme class imbalance, and often lacks multi-modal input. The lightweight genetic algorithm framework is intentionally constrained to ensure practicality on modest hardware, in contrast to computationally intensive neural architecture search. The resulting model achieves near-nnU-Net performance with only a fraction of the cost, a property of direct relevance to real clinical environments. This targeted efficiency and design practicality represent genuine methodological value beyond incremental combination.

The proposed framework’s performance stems from the complementary effect of its components, each contributing distinct advantages:

**Graph-cut preprocessing:** the lightweight graph-cut refinement (implemented using GrabCut with tumor-informed initialization) enhances tumor-background separation during training. Low-grade gliomas often present diffuse boundaries and low contrast in FLAIR MRI, making boundary emphasis essential. The ablation study shows that graph-cut preprocessing provides the largest individual improvement (+3.5% Dice), improving early-stage feature extraction and aiding the U-Net in learning sharper boundaries. Importantly, this step is training-only, adding no runtime cost during inference.

**GA-based architectural refinement:** the constrained genetic algorithm explores only a compact search space tailored to low-resource environments, yet it effectively tunes filter progression in a manner that manually designed models may overlook. Despite the small population size and the limited number of generations (6), the genetic algorithm still contributes a +1.9% Dice improvement and identifies a configuration that generalizes strongly across patients. Unlike full neural architecture search (NAS), which is computationally restrictive, this small-scale genetic algorithm search completes in 17.3 minutes and is therefore practical even on limited GPU setups.

**Compact and clean U-Net backbone:** with only 8.7 million parameters, the final architecture remains lightweight but performs comparably with significantly more complex designs. The HD95 of 1 mm indicates extremely accurate boundary reconstruction—more precise than nnU-Net’s 2.3 mm—increasing suitability for radiotherapy contouring, surgical navigation, and longitudinal monitoring.

**Performance distribution insight:** the combination of preprocessing and GA-based refinement results in a characteristic metric pattern: high **median Dice** (0.9855), indicating near-perfect performance for most slices, and slightly lower **mean Dice** (0.9132), reflecting expected sensitivity to outliers such as tiny tumors or faint infiltrative margins. This mean-median separation is well understood in clinical imaging literature and does not indicate inconsistency; rather, it highlights the method’s robustness across common cases while reflecting the inherent difficulty of a small subset of challenging slices.

While each individual component of the pipeline (graph-cut refinement, U-Net, and GA-based tuning) has been studied previously, their integration into a single, computationally efficient framework tailored specifically for LGG FLAIR segmentation represents a practical contribution. The value of the method lies not in introducing a new architecture, but in demonstrating that lightweight, well-engineered components can achieve accuracy comparable to large automated systems such as nnU-Net at a fraction of the computational cost.

The performance gain from the manually tuned variant (Dice = 0.8350) to the GA-optimized architecture (Dice = 0.9132) is substantial. This improvement arises primarily from the GA’s ability to identify a non-intuitive progression of filter depths that balances feature extraction capacity with model regularization. Unlike manual tuning where filter sizes typically grow in powers of two, the GA frequently explored asymmetric configurations and ultimately selected an architecture with moderate early-layer capacity and an expanded bottleneck (384 filters). This configuration enhanced the model’s ability to preserve boundary details after Graph-Cut preprocessing, which we confirmed through feature-map inspection during training.

### Comparison with nnU-Net

nnU-Net is widely recognized as one of the strongest baselines for medical image segmentation due to its extensive automatic optimization and heavy data augmentation pipeline. However, this strength comes at a high computational cost: 1000 training epochs, multiple stages of preprocessing, and large batch sizes that require powerful GPUs.

In this controlled comparison, where both methods were trained on the same dataset and patient-level splits:

- The proposed method matches nnU-Net’s peak accuracy (0.9134 EMA Dice)
- The **median Dice** improves dramatically (0.9855 vs. 0.9134), indicating stronger reliability across typical slices
- The **boundary accuracy** (HD95) is more than twice as precise (1.0 mm vs. 2.3 mm)
- The training cost is reduced by **roughly 90%**

These findings demonstrate that a carefully constructed pipeline—balancing classical boundary refinement, targeted architecture search, and efficient CNN modeling can rival nnU-Net in performance while being an order of magnitude more efficient. This indicates that high segmentation quality does not always require heavy NAS pipelines or deep transformer-based backbones.

### Clinical Relevance

The motivation for designing a lightweight pipeline is strongly tied to real clinical constraints. Many radiology departments—especially those outside major research centers—rely primarily on FLAIR imaging for LGG assessment and do not have access to multi-modal MRI sequences. In addition, training or deploying large models such as nnU-Net often exceeds available GPU memory and compute time. Therefore, achieving high segmentation accuracy under strict efficiency constraints is essential for widespread adoption in actual clinical workflows.

The system’s efficiency makes it suitable for real-time or near-real-time clinical workflows:

- Full 3D MRI segmentation in under 15 seconds
- High specificity (0.9997) reduces unnecessary false-positive corrections
- High precision (0.9523) produces clean tumor borders, useful for radiotherapy contouring
- Low HD95 (1 mm) supports integration into surgical planning and navigation systems

These characteristics allow easy deployment on standard clinical hardware, including mid-range GPUs commonly found in radiology departments.

### Limitations

Despite the strong performance achieved by the proposed framework, several important limitations must be acknowledged.

#### Slice-wise 2D segmentation

The model operates on 2D FLAIR slices and therefore does not explicitly exploit volumetric continuity across adjacent MRI slices. While the proposed Graph-Cut–enhanced training improves boundary localization and partially compensates for this limitation, fully 3D or 2.5D architectures could better capture inter-slice contextual information, especially for tumors with complex spatial morphology. Future work will extend the framework toward volumetric segmentation.

#### Single-modality input (FLAIR only)

The system is designed specifically for FLAIR-only imaging to ensure applicability in resource-constrained clinical settings. However, multimodal MRI sequences (T1, T1ce, T2) provide complementary tissue information that could further enhance segmentation robustness. While the current results demonstrate that high accuracy can be achieved using FLAIR alone, multi-modal fusion remains an important direction for future investigation.

#### Dataset size and limited external generalizability

The dataset includes 110 patients, which is moderate for LGG research but still limited for capturing the full variability encountered in large multi-center clinical practice. Furthermore, the test set contains only 12 patients. Although this follows established patient-level evaluation protocols for LGG segmentation, broader validation across institutions, scanners, and acquisition protocols is required before large-scale clinical deployment.

#### Limited genetic algorithm search space

The genetic algorithm was intentionally constrained to a small population size and only six generations to maintain computational efficiency. While this lightweight design enables rapid optimization, it may restrict exploration of a broader architectural space. Nevertheless, the consistent convergence patterns observed across generations (Fig. 3 and Fig. 4) indicate that the search stabilized under these constraints.

#### Graph-Cut preprocessing complexity

The training-only Graph-Cut preprocessing step, although beneficial for boundary enhancement, introduces additional algorithmic components and hyperparameters (e.g., Gaussian mixture modeling and regularization weights) that may require tuning when applied to datasets with different intensity characteristics. While this step adds no inference-time cost, its dataset-dependent nature represents an additional layer of methodological complexity.

#### Lack of external multi-center validation

The current evaluation is limited to a single public dataset. Performance on data acquired across different institutions, scanner vendors, and imaging protocols remains to be established, and constitutes an essential next step for assessing real-world applicability.

## Future Work

This work provides a foundation for several promising research directions. Future extensions will include:

- **Advanced graph-cut formulations:** extending the energy model to automatically adapt pairwise and unary terms
- **3D volumetric variants** of the hybrid pipeline
- **Multi-modal integration** (T1/T1ce/T2 + FLAIR)
- **External, multi-center evaluations** to assess robustness across institutions
- **Uncertainty quantification** for clinical decision support
- **Hybrid CNN-transformer architectures** under the same efficient design philosophy

In summary, this study demonstrates that a strategically designed hybrid pipeline—pairing classical boundary refinement with targeted architectural optimization—can deliver segmentation accuracy comparable to the leading benchmark automated systems like nnU-Net while reducing training cost by nearly 90%. This balance of accuracy, efficiency, and boundary sharpness positions the method as a practical and clinically deployable solution for LGG segmentation in FLAIR MRI.

## Conclusion

This study presents a computationally efficient hybrid framework for LGG brain tumor segmentation in FLAIR MRI, combining graph-cut-based refinement, a lightweight GA-optimized U-Net, and a minimal post-processing pipeline. The method achieves a mean Dice of 0.9132, a median Dice of 0.9855, and an HD95 of 1 mm—demonstrating both strong overall accuracy and high reliability across typical slices. Despite its compact design, the framework matches the performance of nnU-Net while reducing training time by nearly 90% and requiring substantially fewer computational resources. These results highlight that carefully orchestrated preprocessing, targeted architectural tuning, and efficient training protocols can achieve accuracy competitive with widely known systems while remaining practical for clinical deployment on standard hardware.

Importantly, the goal of this study is not to replace large-scale frameworks but to demonstrate that clinically viable performance can be achieved using models that are significantly more compact, faster to train, and easier to deploy. This is particularly relevant for environments where nnU-Net-scale training is impractical.

Future work will explore enhanced graph-cut formulations, multi-modal extensions, 3D volumetric segmentation, external multi-center validation, and hybrid CNN-transformer designs. Overall, this work demonstrates a promising pathway toward clinically practical, resource-efficient tumor segmentation models suitable for real-world radiology settings.

## Data Availability Statement

All data used in this study are publicly available through The Cancer Imaging Archive (TCIA) [10] and are also publicly mirrored on Kaggle. The dataset is fully anonymized and requires no special access permissions.

All code, trained models, and configuration files will be made publicly available upon acceptance of this manuscript at: https://github.com/ReemMomen/medical-image-segmentation. During peer review, the materials are available upon reasonable request from the corresponding author.

## Ethics Statement

This research uses fully anonymized, publicly accessible MRI data from TCIA. No identifiable patient information is included, and according to institutional guidelines, no ethical approval was required.

## Author Contributions

Conceptualization, methodology, validation, investigation: R.M.M., E.M., A.A., H.Z.Z. Software and implementation: R.M.M. Writing–original draft: R.M.M. Writing–review and editing: E.M., A.A., H.Z.Z. Supervision: E.M., A.A., H.Z.Z.

## Acknowledgments

The authors thank the Faculty of Science, Al-Azhar University, for providing computational guidance and academic support throughout this research.

## References

1. Erena Siyoum Biratu, Friedhelm Schwenker, Yehualashet Megersa Ayano, and Taye Girma Debelee. A survey of brain tumor segmentation and classification algorithms. Journal of Imaging, 7(9):179, 2021.

2. Nahian Siddique, Sidike Paheding, Colin P. Elkin, and Vijay Devabhaktuni. U-net and its variants for medical image segmentation: A review of theory and applications. IEEE Access, 9:82031–82057, 2021.

3. Leland S. Hu, Andrea Hawkins-Daarud, Lujia Wang, Jing Li, and Kristin R. Swanson. Imaging of intratumoral heterogeneity in high-grade glioma. Cancer Letters, 477:97–106, 2020.

4. Jun Cheng, Wei Huang, Shilei Cao, Rongchang Yang, Wei Yang, Zhongchao Yun, Zhentai Wang, and Qianjin Feng. Enhanced performance of brain tumor classification via tumor region augmentation and partition. PLOS ONE, 12(10):e0186856, 2017.

5. Olaf Ronneberger, Philipp Fischer, and Thomas Brox. U-net: Convolutional networks for biomedical image segmentation. In MICCAI 2015, pages 234–241, 2015.

6. Zongwei Zhou, Md Mahfuzur Rahman Siddiquee, Nima Tajbakhsh, and Jianming Liang. Unet++: A nested u-net architecture for medical image segmentation. In Deep Learning in Medical Image Analysis and Multimodal Learning for Clinical Decision Support, pages 3–11. Springer, 2018.

7. Gusztáv Gaál, Balázs Maga, and András Lukács. Attention u-net based adversarial architectures for chest x-ray lung segmentation. arXiv preprint arXiv:2003.10304, 2020.

8. Xiaocong Chen, Lina Yao, and Yu Zhang. Residual attention u-net for automated multi-class segmentation of covid-19 chest ct images. arXiv preprint arXiv:2004.05645, 2020.

9. Jie Hu, Li Shen, and Gang Sun. Squeeze-and-excitation networks. In Proceedings of the IEEE/CVF Conference on Computer Vision and Pattern Recognition (CVPR), pages 7132–7141, 2018.

10. Spyridon Bakas et al. Advancing tcga glioma mri collections with expert segmentation labels. Scientific Data, 4(1):1–13, 2017. Dataset mirror on Kaggle.

11. Pengzhen Ren, Yun Xiao, Xiaojun Chang, Po-Yao Huang, Zhihui Li, Bingkuan Chen, Xinchao Wang, and Yi Yang. A comprehensive survey of neural architecture search: Challenges and solutions. ACM Computing Surveys, 54(4):1–34, 2021.

12. Fabian Isensee et al. nnu-net: Self-configuring method for biomedical image segmentation. arXiv:2011.00848, 2020.

13. Yuan-Xing Zhao, Yan-Ming Zhang, and Cheng-Lin Liu. Bag of tricks for 3d mri brain tumor segmentation. In BrainLes 2019, MICCAI, pages 210–220. Springer, 2020.

14. Chen Chen, Xiaopeng Liu, Meng Ding, et al. 3d dilated multi-fiber network for real-time brain tumor segmentation in mri. In MICCAI 2019, pages 184–192, 2019.

15. Hu Cao, Yueyue Wang, Joy Chen, Dongsheng Jiang, Xiaopeng Zhang, Qi Tian, and Manning Wang. Swin-unet: Unet-like pure transformer for medical image segmentation. In European Conference on Computer Vision (ECCV) Workshops, pages 205–218. Springer, 2022.

16. Olivier Petit, Nicolas Thome, Cĺement Rambour, Löıc Themyr, Toby Collins, and Luc Soler. U-net transformer: Self and cross attention for medical image segmentation. In Machine Learning in Medical Imaging, pages 267–276. Springer, 2021.

17. Jyotsna Dogra, Shruti Jain, and Ashutosh Sharma. Brain tumor detection from mr images employing fuzzy graph cut technique. Recent Advances in Computer Science and Communications, 13(3):362–369, 2020.

18. Roaa Soloh, Wael Rhmann, and Alaa Shantaf. Brain tumor segmentation based on *α*-expansion graph cut. Journal of King Saud University - Computer and Information Sciences, 36(2):101954, 2024.

19. Annu Lambora, Kunal Gupta, and Kriti Chopra. Genetic algorithm—a literature review. In 2019 International Conference on Computing, Communication, and Intelligent Systems (ICCCIS), pages 380–384. IEEE, 2019.

20. Mitsuo Gen, Runwei Cheng, and Lin Lin. Genetic algorithms and engineering optimization. Wiley, 2023.

21. Sajid Iqbal, M Usman Ghani, Tanzila Saba, and Amjad Rehman. Brain tumor segmentation in multi-spectral mri using convolutional neural networks (cnn). Microscopy Research and Technique, 81(4):419–427, 2018.

22. Lele Chen, Yue Wu, Adora DSouza, et al. Mri tumor segmentation with densely connected 3d cnn. SPIE Medical Imaging, 10574:357–364, 2018.

23. Naji Mahyoub, James Jun, Islam Reda, Abeer Al-Azazi, and Tyler L. Kline. Brain tumor segmentation in fluid-attenuated inversion recovery mri scans using a residual u-net architecture. International Journal of Advanced Computer Science and Applications, 14(7):1082–1090, 2023.

24. Yudong Wu, Zhihai Zhao, et al. Intelligent segmentation system for mri brain tumor images based on optimized region growing. IEEE Access, 8:13925–13933, 2020.

25. Majid Behzadpour, Ebrahim Azizi, Kai Wu, and Bengie Ortiz. Enhancing brain tumor segmentation using channel attention and transfer learning. arXiv preprint, 2025, 2025.

26. M. Baldeon-Calisto and S. K. Lai-Yuen. Evolutionary neural architecture search for retinal vessel segmentation. IEEE Access, 8:179–192, 2020.

27. Carsten Rother, Vladimir Kolmogorov, and Andrew Blake. Grabcut: Interactive foreground extraction using iterated graph cuts. In ACM Transactions on Graphics (SIGGRAPH), volume 23, pages 309–314. ACM, 2004.

28. Fausto Milletari, Nassir Navab, and Seyed-Ahmad Ahmadi. V-net: Fully convolutional neural networks for volumetric medical image segmentation. In 2016 Fourth International Conference on 3D Vision (3DV), pages 565–571. IEEE, 2016.

29. Agung Setiawan. Image segmentation metrics in skin lesion: accuracy, sensitivity, specificity, dice, iou. CENIM, pages 97–102, 2020.

30. Carole Sudre, Wenqi Li, et al. Generalised dice overlap as a deep learning loss function. In DLMIA and ML-CDS, MICCAI, pages 240–248. 2017.

31. Frank Wilcoxon. Individual comparisons by ranking methods. Biometrics Bulletin, 1(6):80–83, 1945.

32. Robert Rosenthal. Meta-Analytic Procedures for Social Research. Sage Publications, 1991.

